# The East London Parkinson’s Disease Project – A case-control study in a diverse population

**DOI:** 10.1101/2024.11.24.24317730

**Authors:** Alexandra Zirra, Kamalesh C. Dey, Ellen Camboe, Sheena Waters, Tahrina Haque, Brook Huxford, Harneek Chohan, Natalie Donkor, Joshua Kahan, Aaron Ben-Joseph, David A. Gallagher, Caroline Budu, Thomas Boyle, Cristina Simonet, Andrew J Lees, Charles R. Marshall, Alastair J. Noyce

## Abstract

**Background:** There is a relative dearth of research on patients with Parkinson’s disease (PD) from under-represented ethnic groups in the United Kingdom.

**Objectives:** The East London Parkinson Disease project seeks to understand the clinical manifestations and determinants of PD in a diverse population.

**Methods:** Patients with PD were recruited from the Royal London Hospital. Healthy controls came from community engagement events and partners of patients. Data on clinical features assessed by motor and non-motor scales were collected between January 2019 and February 2024, and compared between groups. Parametric, non-parametric tests, and unmatched logistic models, adjusted for age, gender and duration of disease were used.

**Results:** We assessed 218 patients with PD and 90 controls. Among them, 50% of patients and 64% controls identified as South Asian or Black. Males comprised 63% of patients and 70% of controls. After adjusting for age, gender, disease duration and treatment burden, South Asian and Black patients had significantly worse motor scores compared to White patients (mean [SD], 42.2 [18.8], and 47 [16.6] vs 35.2 [16.4], p<0.001 and p<0.001). Cognitive impairment was more prevalent in South Asian (73%) and Black patients (75%) than in White patients (45%, p=0.002).

**Conclusions:** Our results suggest that patients with PD from South Asian and Black ethnic groups may have more severe motor and certain non-motor features, including cognitive impairment, compared to White patients.

## Introduction

Although progress in understanding the clinical types and patterns of Parkinson’s disease (PD) progression have been made using clinical, genetic and transcriptomic data^<u>1,2,3</u>^, there remains a lack of research in diverse populations^<u>4</u>^. Most studies have focused predominantly on White, relatively affluent, well-educated patients, who attend tertiary neurology services^<u>5</u>^. Even landmark initiatives seeking to develop markers of PD progression, such as the Parkinson’s Progression Marker Initiative (PPMI) or Parkinson’s Disease Biomarkers Program (PDBP), have enrolled more than 95% White participants^<u>6,7</u>^.

Several large initiatives aimed at population-specific risk factors for Parkinson’s and PD progression, such as LARGE-PD^<u>8</u>^, BLAAC PD^<u>9</u>^, and GP2^<u>5</u>^, have started to address diversity in PD research. However, these are predominantly focused on understanding the genetic basis of PD risk.

The East London Parkinson’s Disease (ELPD) project was established to carry out research in a highly diverse population from East London with free access to a publicly-funded, free at the point of service healthcare system (namely the National Health Service - NHS). We aimed to recruit participants so that our sample reflected the underlying population structure in East London, where >40% identify as South Asian and 7% as Black, and ∼45% are from the lowest UK deprivation group^<u>10</u>^. The focus of present work was to describe the clinical features by ethnicity in patients and controls recruited to the ELPD project.

## Methods

### Study design and participants

Research Ethics Board approval was received on 29th November 2018 from the South West - Central Bristol Research Ethics Committee, under the reference 18/SW/0255, IRAS ID 242395. A register of patients with PD and parkinsonism was created locally at the Barts Health NHS Trust. This was then used to recruit patients from the Movement Disorder outpatient clinic at the Royal London Hospital.

The inclusion criteria for the patient group were as follows: patients over the age of 18, with a clinical diagnosis of Parkinson’s, able to consent or have appropriate next-of-kin/proxy for consent. The clinical diagnosis of PD was made by movement disorder consultants according to MDS 2015 criteria^<u>11</u>^. The exclusion criteria for the patient group included: secondary parkinsonism (such as vascular or drug-induced parkinsonism), alternative neurological or psychiatric diagnoses (other movement disorders, including stroke and motor neurone disease, and unrelated dementia). Controls were recruited through several approaches: spouses of patients, people attending outpatient clinics for indications other than neurological symptoms, as well as public involvement events. Inclusion criteria were: age above 35 years, absence of parkinsonism, ability to consent. Exclusion criteria were neurological or psychiatric diagnoses with the exception of idiopathic intracranial hypertension.

Enrolment strategies were focused on increasing participation from under-represented populations, such as participants from South Asian and Black ethnic backgrounds, and reducing barriers to research. To achieve this, we recruited a diverse, multi-lingual team with researchers that represented the same communities we sought to enrol in the study. We offered home visits, at varied times of the day, as an alternative to scheduled clinic-based visits. We translated relevant study materials (information sheets, consent forms and certain scales). Patient and public involvement events were organised to increase awareness of PD in the East London community, keep research participants up-to-date on study progress and aid recruitment of healthy controls.

### Data collection

Data collection for the study started in January 2019. Study visits were undertaken at the Royal London Hospital, or in the participant’s home.

The study protocol consists of one mandatory clinical visit for both patients and controls (Figure 1, Supplementary Figure 1), and second and third optional visits for patients (Supplementary Figure 2). Demographic data, clinical motor and non-motor assessments (e.g. hearing and vision performance captured by computerised psychophysical tests), and biological samples for biomarkers and genetic analysis of PD were collected (Figure 1). The participant postcode was used to derive the IMD - Index of Multiple Deprivation^<u>12</u>^, with 1 representing most deprived, and 10 least deprived decile.

**Figure 1.**
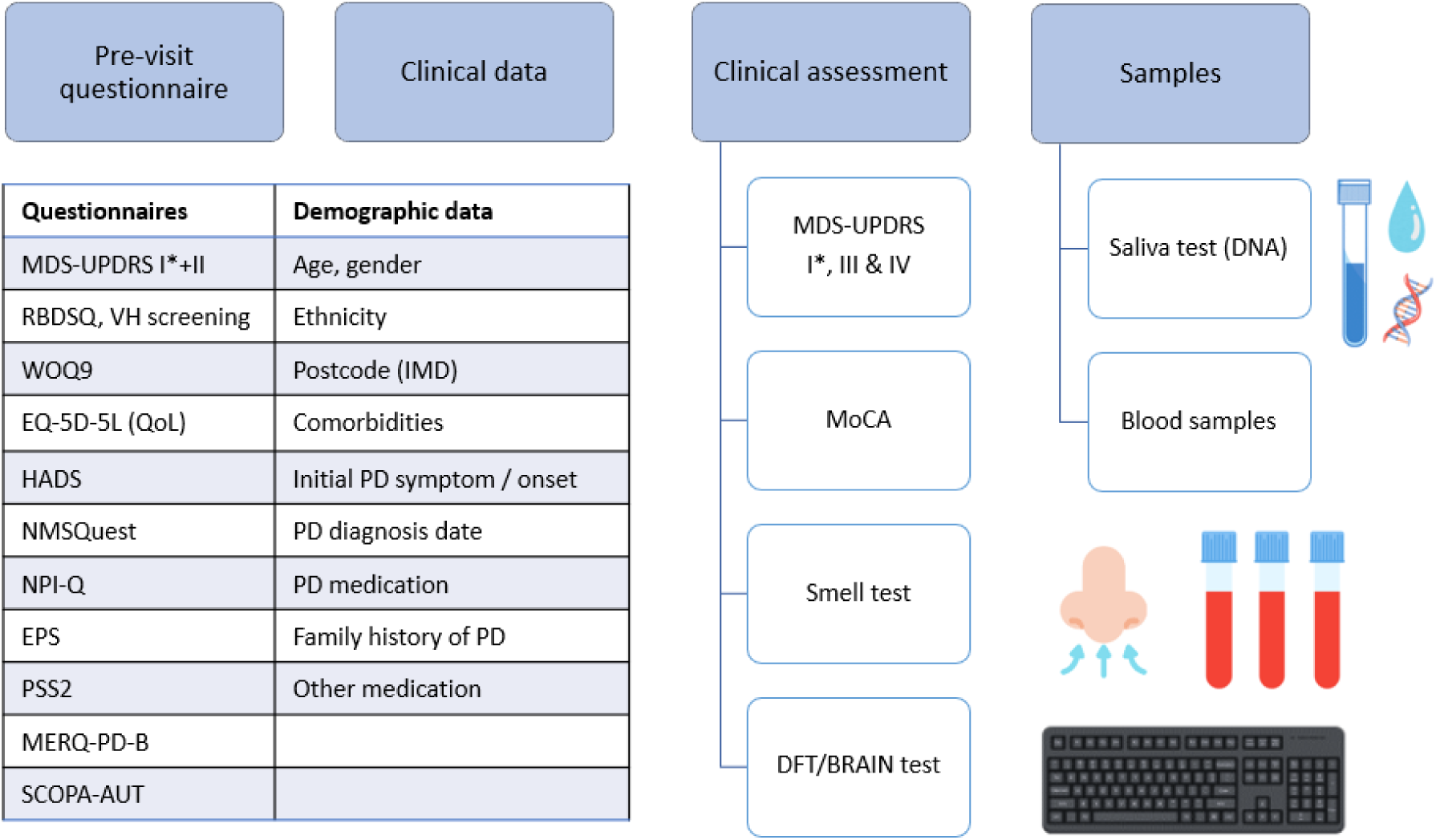
Mandatory visit for the East London Parkinson’s Disease study. MDS-UPDRS - MDS-Unified Parkinson’s Disease Rating Scale, MoCA - Montreal Cognitive Assessment, RBDSQ - REM sleep disease screening questionnaire, VH - visual hallucinations, WOQ9 - Wearing OFF questionnaire, EQ-5D-5L QoL - Quality of Life Questionnaire, HADS - Hospital Anxiety and Depression Questionnaire, NMSQ - Non-Motor Symptoms Questionnaire, EPS - Epworth Sleepiness Questionnaire, PSS2 - Parkinson’s Sleep Scale 2, MERQ-PD-B - Mini-environmental Risk factor Questionnaire in Parkinson’s Disease version B, SCOPA-AUT - Scales for Outcomes in Parkinson’s Disease - Autonomic Dysfunction, PD - Parkinson’s disease. Created with Canva^<u>48</u>^.

At the mandatory clinical visit, the following assessments & questionnaires were used: motor symptoms (MDS-UPDRS - Movement Disorders Society - Unified Parkinson’s Disease Rating Scale^<u>13</u>^, BRAIN test - BRadykinesia Akinesia INcoordination test^<u>14</u>^, DFT - Distal Finger Tapping test^<u>15</u>^), non-motor symptoms (NMSQ - Non-Motor Symptoms Questionnaire^<u>16</u>^, Epworth Sleepiness Questionnaire^<u>17</u>^, PSS2 - Parkinson’s Sleep Scale 2^<u>18,19</u>^, RBDSQ - REM-sleep behaviour disease screening questionnaire^<u>20</u>^, an abbreviated 6-item version of the UPSIT - University of Pennsylvania Smell Identification Test^<u>21</u>^), cognitive symptoms (MoCA - Montreal Cognitive Assessment^<u>22</u>^), quality-of-life (EQ-5D-5L Quality-of-Life Questionnaire^<u>23</u>^), psychiatric symptoms (HADS - Hospital Anxiety and Depression Questionnaire^<u>24</u>^), risk factors (MERQ-PD- B - Mini-environmental Risk factor Questionnaire in Parkinson’s Disease version B^<u>25</u>^). For the BRAIN test, the following were described: KS - kinesia score, number of taps in 30 seconds, AT - akinesia time, mean dwell-time on each key, IS - incoordination score,variance of travelling time between key taps^<u>26</u>^. The hemibody with the higher MDS-UPDRS III score for patients, and the lowest KS for controls, were used.

Pseudo-anonymised data were stored securely on Queen Mary University of London servers for analysis and are available upon request.

### Data analysis and statistical methods

Data were recorded in Microsoft Access database format and exported as ‘CSV’ files. We defined normal cognition (MoCA>25), mild cognitive impairment (MoCA=19-25), and dementia (MoCA<19)^<u>22</u>^. This was either administered in English or a validated Bengali translation by our team’s Bengali-speaking team members (see Appendix 1).

Statistical analysis and figure generation were completed with Python 3.10.5 in Jupyter Notebook 6.4.8. Statistical analysis included T-test, Mann-Whitney U, Χ^2^ and Fisher’s exact tests, for continuous and categorical values. Logistic regression was adjusted for age, gender and time from onset. The MDS-UPDRS III, was also adjusted for levodopa-equivalent daily dose. Bonferroni correction for multiple comparisons was used as follows: 3 ethnic groups (White, South Asian, and Black) & 4 main analyses (MDS-UPDRS III, IV, MoCA and NMSQ), significance cut-off p<0.004.

### Data sharing

The code for analysis will be publicly available on GitHub. The raw data will be made available by request.

## Results

Since 2018, a total of 218 patients and 90 controls have been recruited to the ELPD project. Of patients, 85 (39%), and of the controls, 56 (62%) identified as South Asian (Table 1). Males represented 137 (63%) and 63 (70%) of healthy controls. The age at assessment was higher in patients than controls (mean [SD], 68.5 [10.8] years vs 62.7 [10.9], p<0.001). Our patients had an index of multiple deprivation decile of 3 [1] (median [IQR]) consistent with being in the most deprived tertile in the UK.

**Table 1.** Demographics and clinical data - Patients with Parkinson’s disease (PwP) and healthy controls (HC). a - Kruskall Wallis test; b - Chi-squared test; c - Fisher’s exact test. IMD - Index of Multiple Deprivation; SD - standard deviation; IQR - interquartile range; bold - p < 0.004 (Bonferroni adjusted for multiple comparisons)

| Whole cohort |  | PwP | HC | p, t(dof) |
| --- | --- | --- | --- | --- |
|  |  | n = 218 | n = 90 |  |
| **Age at assessment (mean ± SD)** | | 68.5 ± 10.8 | 62.7 ± 10.9 | **<0.001<sup>e</sup>, 12873** |
| **Male (n, %)** | All (n, %) | 137, 63% | 63, 70% | 0.287<sup>b</sup>, 1.136(1) |
| **Ethnicity (n, %)** | White (n, %) | 96, 44% | 30, 33 % | **<0.001<sup>c</sup>, 14.79(2)** |
|  | South Asian (n, %) | 85, 39% | 56, 62 % |  |
|  | Black (n, %) | 24, 11% | 2, 2 % |  |
| **Comorbidities** | HTN (n, %) | 112, 52% | 30, 42% | 0.174<sup>b</sup>, 0.049(1) |
|  | T2DM (n, %) | 68, 31% | 21, 29% | 0.825<sup>b</sup>, 0.049(1) |
|  | Smoking (n, %) | 61, 29% | 2, 13% | 0.197<sup>b</sup>, 1.67(1) |
| **Time from diagnosis, years (median ± IQR)** | | 4.8 ± 6.9 | - | - |
| **Duration of symptoms on assessment, years (median ± IQR)** | | 6.3 ± 7.4 | - | - |
| **Age at diagnosis, years (mean ± SD)** | | 61.3 ± 12 | - | - |
| **Age at symptom onset, years (mean ± SD)** | | 61 ± 12 | - | - |
| **LEDD (median ± IQR)** | | 510 ± 525 | - | - |
| **UPDRS I (mean ± SD)** | | 16.2 ± 8.8 | - | - |
| **UPDRS II (mean ± SD)** | | 19.5 ± 11 | - | - |
| **UPDRS III (mean ± SD)** | | 38.6 ± 18.1 | - | - |
| **UPDRS IV (mean ± SD)** | | 4.7 ± 4.3 | - | - |
| **NMSQ (median ± IQR)** | | 12 ± 10 | - | - |
| **MoCA (mean ± SD)** | | 23.8 ± 4.6 | 25.6 ± 3.4 | 0.024<sup>a</sup>, 5.096 |
| **Smell test (median ± IQR)** | | 2 ± 2 | 3 ± 2 | 0.007<sup>e</sup>, 964 |
| **HADS Depression (mean ± SD)** | | 7.1 ± 4.5 | - | - |
| **HADS Anxiety (mean ± SD)** | | 6.9 ± 4.6 | - | - |
| **RBDSQ (mean ± SD)** | | 4.6 ± 3.1 | - | - |
| **PSS2 (mean ± SD)** | | 19.5 ± 11.3 | - | - |
| **ESS (mean ± SD)** | | 9.4 ± 6.4 | - | - |
| **SCOPA-AUT (mean ± SD)** | | 17.5 ± 10.3 | - | - |
| **EQ5D5L VAS (mean ± SD)** | | 59.4 ± 20.8 | - | - |
| **EQ5D5L index (mean ± SD)** | | 0.47 ± 0.3 | - | - |
| **IMD decile (median ± IQR)** | | 3 ± 1 | - | - |

### Demographic characteristics

Demographic and disease-related data are presented in Table 1. Age at assessment was higher in Black and White compared to South Asian patients (mean [SD], 71.1 [10], and 70.2 [9.5] vs 66.4 [11.4] years), Table 2. Time from diagnosis was 4.8 [6.9] (median, [IQR]) years, and duration of PD symptoms was 6.3 [7.4] years. There were no statistical differences between time from diagnosis and duration of symptoms across ethnic groups.

**Table 2.** Clinical characteristics of patients with Parkinson’s disease in the East London Parkinson’s Disease project. a - Kruskall Wallis test; b - Chi-squared test; c - Fisher’s exact test; d - two-sided T-test; e - Mann-Whitney U test; f - Logistic regression, adjusted for disease duration; g - Logistic regression, adjusted for disease duration, age and gender; h - Logistic regression, adjusted for disease duration, age, gender, and ON-OFF status, j - Logistic regression, adjusted for disease duration, age, gender, and LEDD. SD - standard deviation; IQR - interquartile range; t - T statistic; dof - degrees of freedom; bold - p<0.004 (Bonferroni adjusted for multiple comparisons). Ages and durations reported in years.

|  | White<br>n=96 | South<br>Asian n=85 | Black n=24 | White vs South Asian |  | White vs Black |  | Black vs South Asian |  |
| --- | --- | --- | --- | --- | --- | --- | --- | --- | --- |
|  |  |  |  | Uni | Multi | Uni | Multi | Uni | Multi |
|  |  |  |  | p, t(dof) | p | p, t | p | p, t(dof) | p |
| Age at assessment (mean ± SD) | 70.2 ± 9.5 | 66.4 ± 11.4 | 71.1 ± 10 | 0.015<sup>d</sup>,<br>2.45(179) | - | 0.676<sup>d</sup>, -<br>0.42(118) | - | 0.068<sup>d</sup>, 1.84(107) | - |
| Age at diagnosis (mean ± SD) | 64.5 ± 11 | 61 ± 11.5 | 65.6 ± 11.4 | 0.036<sup>e</sup>, 4818 | - | 0.643<sup>e</sup>, 1081 | - | 0.089<sup>d</sup>, -1.72(107) | - |
| Age at symptom onset (mean ± SD) | 62.6 ± 11.3 | 59.2 ± 12.2 | 64.4 ± 11.9 | 0.055<sup>e</sup>, 4756 | - | 0.505<sup>e</sup>, 1050 | - | 0.068<sup>d</sup>, 1.84(107) | - |
| Time from diagnosis (median ± IQR) | 4.1 ± 6.6 | 5 ± 7.7 | 5.6 ± 4.5 | 0.737<sup>e</sup>,<br>3961.5 | - | 0.570<sup>e</sup>, 1065 | - | 0.832<sup>e</sup>, 1049.5 | - |
| Duration of symptoms on assessment (median ± IQR) | 6 ± 7.6 | 6.8 ± 7.3 | 6.5 ± 3.7 | 0.952<sup>e</sup>, 4101 | - | 0.945<sup>e</sup>, 1141 | - | 0.939<sup>e</sup>, 1009 | - |
| Duration of symptoms at diagnosis (median ± IQR) | 1.2 ± 1.5 | 1 ± 1.9 | 1 ± 0.9 | 0.171<sup>e</sup>,<br>4561.5 | - | 0.247<sup>e</sup>, 1329 | - | 0.968<sup>e</sup>, 1026 | - |
| LEDD, mg (median ± IQR) | 481 ± 503 | 625 ± 535 | 510 ± 363 | 0.286<sup>e</sup>, 3705 | - | 0.942<sup>e</sup>, 1140.5 | - | 0.390<sup>e</sup>, 902 | - |
| UPDRS I (mean ± SD) | 14.6 ± 7.7 | 18.7 ± 9.6 | 15.4 ± 7.8 | 0.004<sup>a</sup>, 8.436 | 0.067<sup>f</sup>, **<0.001<sup>g</sup>** | 0.635<sup>e</sup>, 968 | 0.036<sup>f</sup>, 0.693<sup>g</sup> | 0.137<sup>d</sup>, -1.5(99) | 0.004<sup>f</sup>, 0.068<sup>g</sup> |
| UPDRS II (mean ± SD) | 16.1 ± 9.2 | 23.5 ± 11.7 | 21.8 ± 9.5 | **<0.001<sup>a</sup>**, 18.2 | **<0.001<sup>f</sup>**, **<0.001<sup>g</sup>** | 0.012<sup>e</sup>, 704.5 | 0.962<sup>f</sup>, 0.005<sup>g</sup> | 0.519<sup>d</sup>, -0.64(103) | 0.013<sup>f</sup>, 0.356<sup>g</sup> |
| UPDRS III (mean ± SD) | 35.2 ± 16.4 | 42.2 ± 18.8 | 47 ± 16.6 | 0.013<sup>e</sup>, 2797 | 0.148<sup>f</sup>, **<0.001<sup>g</sup>**,<br>**<0.001<sup>h</sup>**, **<0.001<sup>j</sup>** | **0.002<sup>e</sup>**, 650.5 | 0.727<sup>f</sup>, **0.001<sup>g</sup>**,<br>**<0.001<sup>h</sup>**, **<0.001<sup>j</sup>** | 0.266<sup>d</sup>, 1.12(100) | 0.157<sup>f</sup>, 0.473<sup>g</sup>,<br>0.376<sup>h</sup>, 0.374<sup>j</sup> |
| UPDRS IV (mean ± SD) | 3.5 ± 3.7 | 4.8 ± 4.8 | 6 ± 3.8 | 0.11<sup>a</sup>, 2.554 | 0.078<sup>f</sup>, 0.033<sup>g</sup>,<br>0.031<sup>j</sup> | 0.006<sup>e</sup>, 643 | 0.009<sup>f</sup>, **0.002<sup>g</sup>**,<br>**<0.001<sup>j</sup>** | 0.160<sup>e</sup>, 1055 | 0.403<sup>f</sup>, 0.189<sup>g</sup>,<br>0.093<sup>j</sup> |
| NMSQ (median ± IQR) | 11 ± 8 | 13.5 ± 11.2 | 12 ± 9 | 0.20<sup>a</sup>, 5.43 | 0.103<sup>f</sup>, **0.002<sup>g</sup>** | 0.532<sup>e</sup>, 881 | 0.045<sup>f</sup>, 0.399<sup>g</sup> | 0.438<sup>e</sup>, 747 | 0.009<sup>f</sup>, 0.279<sup>g</sup> |
| MoCA (mean ± SD) | 24.9 ± 4.2 | 22.2 ± 4.7 | 22.4 ± 4.8 | **<0.001<sup>e</sup>**,<br>**3235** | 0.027<sup>f</sup>, 0.481<sup>g</sup> | 0.038<sup>e</sup>, 965 | **<0.001<sup>f</sup>**, 0.277<sup>g</sup> | 0.857<sup>d</sup>, 0.181(66) | 0.01<sup>f</sup>, 0.530<sup>g</sup> |

The index of multiple deprivation decile was higher for White than for South Asian, and Black patients (median [IQR], 4 [2], vs 3 [1] and 3 [2], p=0.003), Supplementary Table 2. Hypertension was present more frequently in the patients identifying as Black (75%) compared to South Asian (52%) and White (49%), p=0.069. Type 2 diabetes was more common in patients identifying as South Asian (46%), compared to Black (26%) and White (24%), p=0.006.

Smoking and alcohol consumption were significantly increased in the White patient group (Table 2, Supplementary Table 1). Caffeine consumption, pesticide exposure, and head injury were very similar between groups (Supplementary Table 1).

### Clinical characteristics

There was weak evidence of an older age at symptom onset in White and Black patients, compared to South Asian patients (mean [SD], 62.6 [11.3], and 64.4 [11.9] vs 59.2 [11.3] years, p=0.055, p=0.068 respectively). Similarly, age at diagnosis did not achieve significance, but was lower in the South Asian patients (Table 2).

Time from symptom onset to diagnosis was similar in all three patient groups - White versus South Asian patients, and versus Black patients (median [IQR], 1.2 [1.5], vs 1 [1.9] and 1 [0.9] years, p=0.247). Symptom duration on assessment in South Asian, Black were similar to White patients (median [IQR], 6.8 [7.3], 6.5 [3.7] vs 6 [7.6] years, p=0.973 and p=0.898). There was no evidence that median levodopa equivalent daily doses (LEDD) were higher in the South Asian group than White and Black patients (median [IQR], 625 [535], 481 [503], vs 510 [363], p=0.286 and p=0.390 respectively).

MDS-UPDRS III motor scores were significantly higher in the South Asian and Black groups compared to White patients (mean [SD], 42.2 [18.8], 47 [16.6], vs 35.2 [16.4], p<0.001 and p<0.001 respectively). The magnitude of these differences remained when adjusted for age, gender, disease duration and LEDD. Fewer South Asian patients (10.4%) reported experiencing an ‘OFF’ period during the assessment, compared to White (23%) and Black groups (29.2%), but this was only nominally significant p=0.043. The difference in MDS-UPDRS III remained significant between ethnicities when adjusting for ON-OFF status (p<0.001). Motor complications were also found to be more severe, with MDS-UPDRS IV for the Black patients compared to the White groups (mean [SD], 6 [3.8] vs 3.5 [3.7], p<0.001), Figure 2.

**Figure 2.**
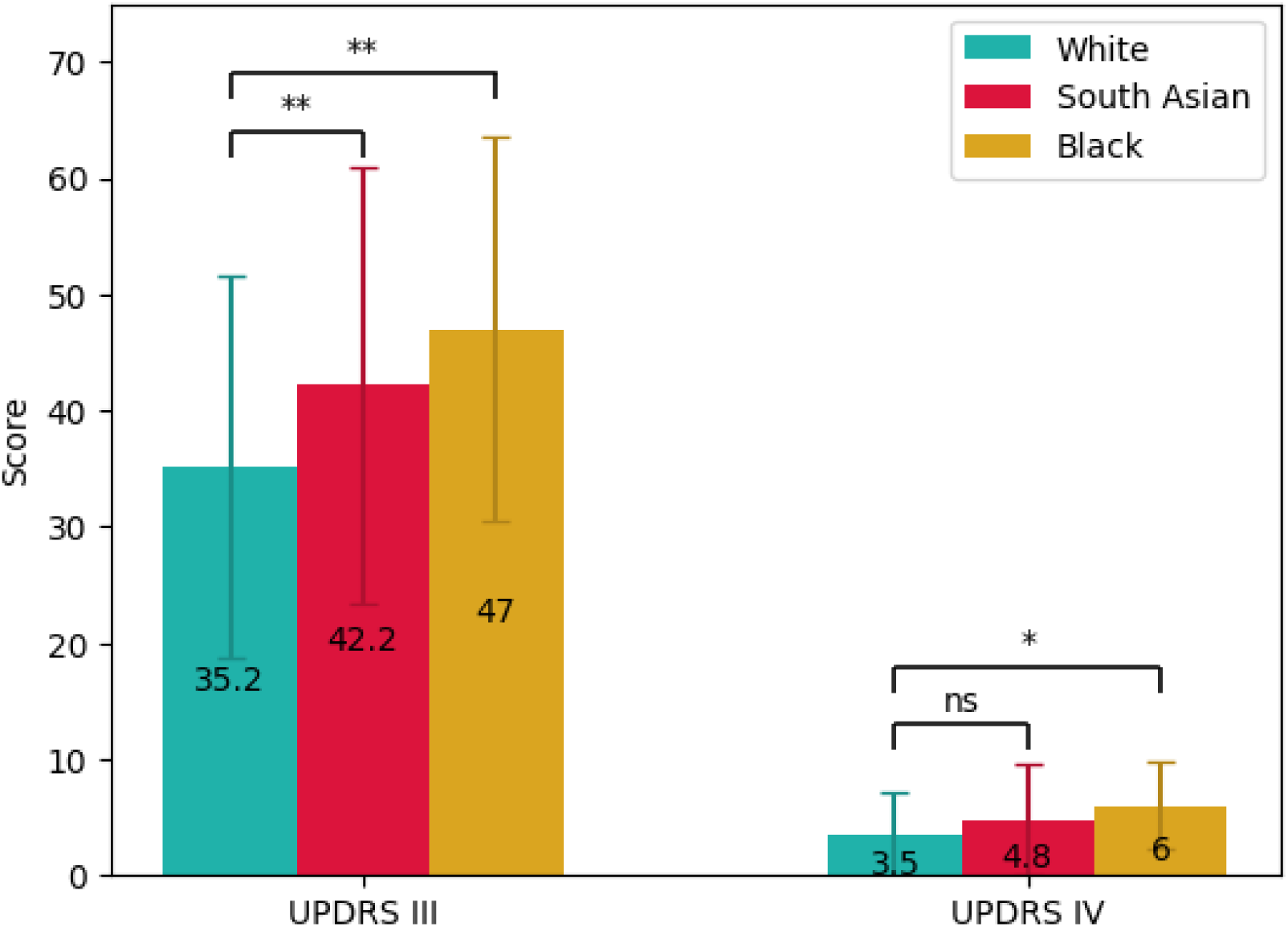
MDS-UPDRS III and IV scores for patients by ethnicity (mean and standard deviation). Ethnicity represented by colour blue (White participants), red (South Asian participants), yellow (Black participants), * - p < 0.004, ** - p < 0.001 (Bonferroni-adjusted for multiple comparisons).

Objective motor assessments showed worse scores for ethnically under-represented patients, with akinesia time (AT) significantly slower in the South Asian compared to the White group (mean [SD], 204.9 [97] vs 153.8 [70.2] ms, p<0.001) when adjusted for age, gender, duration of disease, and ON-OFF state (Supplementary Table 4).

Non-motor and motor experiences of daily living are reflected in the MDS-UPDRS I and II (Table 2). Non-motor symptoms, measured by the NMSQ, were reported more frequently in South Asian compared to White patients (median [IQR], 13.5 [11.2] vs 11 [18], p=0.002).

### Cognitive impairment in ELPD

From 218 patients, 91 White, 52 South Asian and 15 Black patients had reliable cognitive scores on the MoCA (n=158). From 90 controls, only 39 had reliable MoCA scores. For details of participants excluded from this analysis see Appendix 3. There was weak evidence for MoCA scores being higher in controls than patients (mean [SD], 25.6 [3.4] vs 23.8 [4.6], p=0.024). MoCA scores were lower in patients from South Asian and Black ethnicities, compared to the White patients (mean [SD], 22.2 [4.7], 22.4 [4.8] vs 24.9 [4.2], p<0.001 and p<0.001 respectively, Table 2). Based on MoCA scores alone, patients from South Asian and Black groups were identified as having more cognitive impairment than in White groups (38, 73%, and 12, 75%, vs 41, 45%, p=0.002), Figure 3 & Supplementary Table 3.

**Figure 3.**
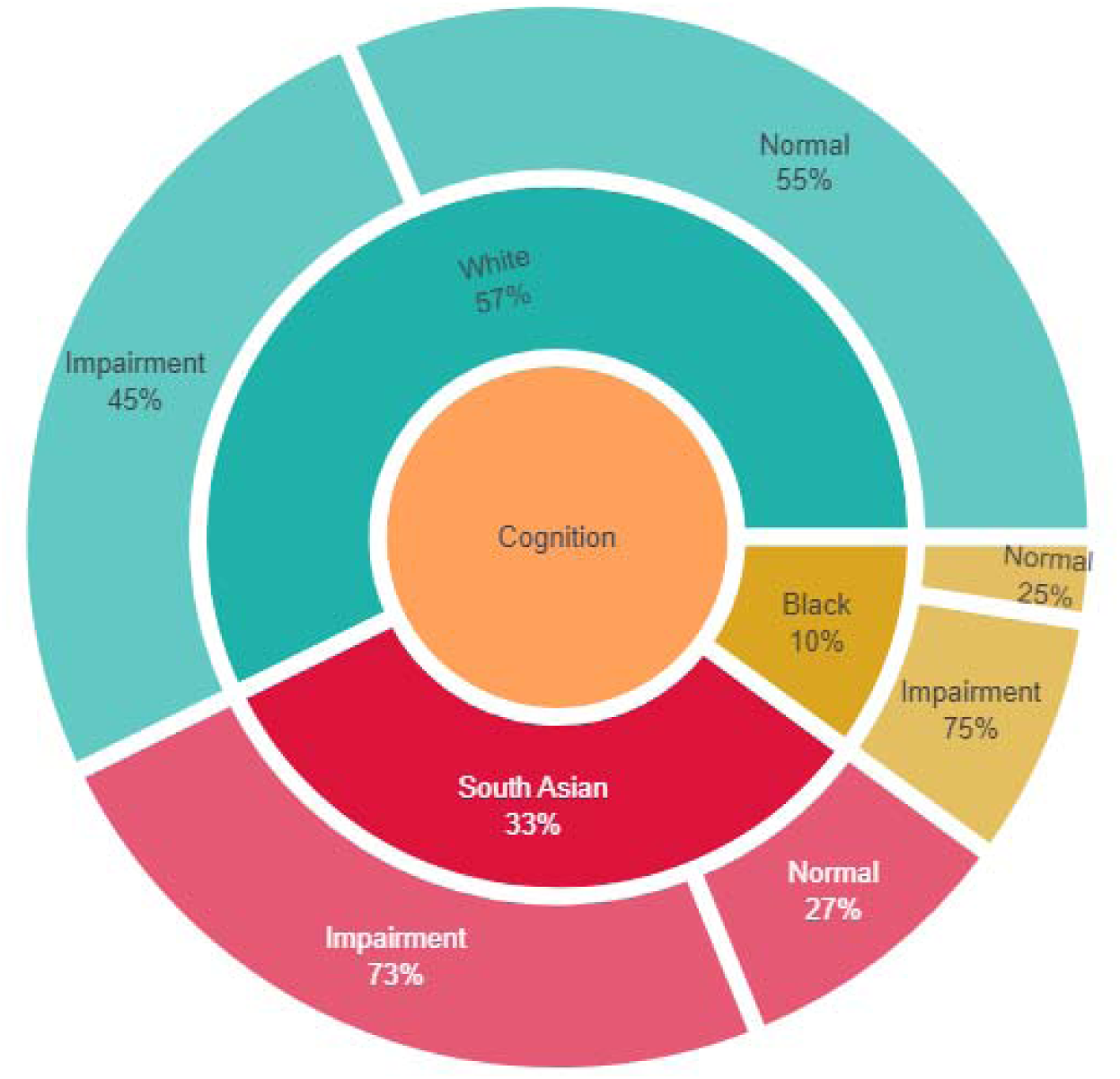
Cognition by ethnicity. Percentage of patients from White (blue), South Asian (red) and Black ethnicity (yellow). Classification based on MoCA: cognitive impairment (<=25), normal cognition (>25).

### Other clinical characteristics

Using a brief 6-item version of the UPSIT, controls had higher scores compared to patients (median [IQR], 3 [2] vs 2 [2], p=0.007). South Asian and White patients had similar smell scores (median [IQR], 2 [3] vs 2 [1], p=0.911), whereas Black patients (1±1) had worse scores (median [IQR], 1 [1], p=0.002, and p=0.037). Patients from South Asian backgrounds had worse scores compared to White for: depression (HADS-D mean [SD], 8.5 [5.2] vs 6.3 [3.8], p=0.019), sleep quality (PSS2 mean [SD] 23.1 [11.7] vs 17.1 [10.2], p=0.008), quality of life index (EQ5D5L index mean [SD] 0.38 [0.31] vs 0.53 [0.26], p<0.001) (see Supplementary Table 2). Black patients had similar scores compared to White for: depression, sleep quality, and quality of life index. No statistical differences between all three ethnicities were seen in the daytime somnolence scores (ESS), autonomic dysfunction (SCOPA-AUT), anxiety (HADS-A) and REM sleep behaviour clinical suspicion (RBDSQ) (Supplementary Table 2).

## Discussion

Here we describe the baseline data in a diverse sample of patients residing in East London receiving ‘free at the point of service’ care via the NHS. Using inclusive recruitment methods, more than 50% of patients and 65% of the control group came from South Asian and Black ethnic groups. We found that motor, non-motor and cognitive features differed by ethnicity, with evidence overall for a more severe burden of disease in under-represented patient groups. There were no differences in time to diagnosis, disease and symptom duration, or treatment differences to account for the worse motor phenotype in South Asian and Black groups.

Our data suggest that South Asian and Black patients had worse motor scores, when adjusting for age, gender, disease duration, and LEDD. We investigated whether assessing patients in ON versus OFF states may be driving the difference in scores. However, MDS-UPDRS III scores were still significantly higher in Black and South Asian patients, when adjusting for this. This is in keeping with other studies that have also shown MDS-UPDRS III reflects disease progression regardless of ON/OFF states^<u>27</u>^. Similarly, motor fluctuations were worse in Black patients compared to the White group, when adjusting for age, gender, disease duration, and LEDD.

The severity of motor scores suggests a requirement for higher doses of medication, or a heavier burden of disease in under-represented patient groups. This may hold true especially in the South Asian patients, as their age at onset was significantly younger. Worse motor severity and fluctuations could be explained by genetic factors, with more atypical features having been reported in underrepresented communities, such as UK-based South Asian and Afro-Caribbean populations^<u>28</u>^. Factors, such as concomitant diabetes, hypertension, or other cardiovascular risk factors could also be an explanation^<u>28</u>^, as well as language/cultural barriers in accurately ascertaining motor fluctuations. In keeping with worse motor symptoms, the subjective assessment of motor activities of daily living (MDS-UPDRS II) in patients from South Asian groups were also found to be more severe compared to White patients.

An objective motor assessment, the BRAIN test, only showed a worse dwell time (AT) when pressing keys on a computer keyboard in South Asian patients. Both our HCs and patients had worse KS compared to other publications^<u>26,29</u>^. This finding, alongside low computer literacy, could explain the less pronounced differences between the groups. More work is needed to understand if the BRAIN test can be used in diverse populations.

There is significant interest in whether the prevalence of dementia/cognitive impairment differs in patients from different ethnic groups^<u>30</u>^. The results from the current study suggest higher rates of cognitive impairment in the Black and South Asian groups compared to White patients. However, the MoCA^<u>22</u>^ was developed for use in White populations and it has been shown to have language, literacy and cultural biases^<u>31,32</u>^. MoCA was shown to perform better when translated and culturally adapted^<u>33</u>^. We used the validated Bengali MoCA for best outcomes. Therefore, we are confident that our study did correctly identify worse cognitive scores in under-represented patients. However, the threshold for defining cognitive impairment required adjustments in translated versions, compared to White population groups^<u>33,32,34</u>^, with lower cut-offs being proposed for older age and/or lower education groups^<u>35</u>^. Our findings confirm that screening tests developed in White, English-speaking countries, may not be the most appropriate tools to investigate cognitive impairment in diverse populations.

Although there were no significant differences between non-motor scores in all three ethnicities, as reported by the NMSQ, we did find significantly worse depression (HADS-D), sleep quality (PSS) in South Asian patients, as well as diminished quality of life measure (EQ5D5L) in both South Asian and Black patients. We also found that patients from South Asian ethnic groups had a slightly higher MDS-UPDRS II score, reflective of a higher non-motor burden in this group.

Non-motor symptoms have been documented to differ in various regions around the globe, with more frequent symptoms present in certain populations. A systematic review from 2020 showed that gastrointestinal symptoms are more prevalent in the East Asian population, whilst depression is worse in East Asian patients^<u>30</u>^. This was in contrast to our data, where we only found that depression, sleep quality and quality-of-life were worse in South Asian patients. However, the systematic review included only two mono-ethnic Indian studies, whereas our study was a head-to-head comparison of the three ethnic groups. Interestingly, a cross-sectional analysis between continents showed a lower burden of non-motor symptoms in Asian patients, especially related to sleep and sexual activity^<u>36</u>^. However, this cross-sectional analysis included both South and East Asian ethnicities in the same category, with White participants having a longer duration of disease. Our study had a consistent methodology, and better balanced clinical characteristics of PD in all three ethnic groups. This disparity between studies could be explained by cultural differences with fewer primary care consultations in under-represented groups, as explored in Simonet et al. in UK-based populations^<u>37,38</u>^. Fortunately, in East London, similar times to diagnosis and levels of treatment between ethnicities suggest an equality of access to primary care, and an unbiased standard of care in secondary settings.

South Asian patients may have a younger age at diagnosis. Large international studies including > 90% White European participants show that age at onset in the White patient group is 60 years (COURAGE-PD, 23andMe, IPDGC)^<u>39,40</u>^. Little is known about the age at onset in South Asian countries^<u>41</u>^, with one study reporting median age at onset as 54 years in Pakistan^<u>42</u>^. Unfortunately, there is relatively low confidence in the reported age of the participants from Bangladesh^<u>43</u>^. In 2020, only 67% of household members were holding birth certificates, with only 54% of the female birth certificates being validated in Bangladesh^<u>44</u>^. However, the age at onset/diagnosis would need further investigation with dedicated prospective incidence studies. Reassuringly, symptom duration at diagnosis in all three major ethnic groups (White, South Asian and Black) was similar, suggesting that in East London, there is an equivalent awareness of PD symptoms in the community and in primary care. This was a positive finding, as other studies show delays in diagnosis and more advanced disease at the time of diagnosis for under-represented patients^<u>45,46,37</u>^. Other studies have shown a higher prevalence in PD in White populations compared to Asian or Black populations^<u>45,46</u>^.

One limitation of our study is that this is a case-control study, therefore it may be affected by sampling bias. We addressed this by running adjusted logistic analyses. A second inherent limitation was the recall bias of patients for their first symptoms and age at diagnosis. However, in most cases, we were able to corroborate with GP records of ‘tremor’, or ‘gait disturbance’, or other movement disorder symptoms. A third limitation in our study is the possibility of diagnostic error. However, as seen from previous studies,^<u>47,11</u>^ diagnostic errors in a specialist movement disorder service are lower than in non-specialist centres. In addition, our patient cohort was around the 5-year mark from diagnosis, therefore less likely to experience a diagnostic error.

Another limitation in our study was the use of different raters over time, which may have introduced inter-rater variability. We mitigated this by undertaking the same clinical training for MDS-UPDRS, MoCA and attending the same movement disorders clinics. Sampling bias and lack of confidence in reported age may be partly responsible for the younger age at assessment for the South Asian group. Despite this, age was not a relevant covariate in most of the statistical models. Future work in our cohort is needed. A final limitation, when analysing the MoCA data, was that we have not used a gold standard to define cognitive impairment in patients with PDD. Steps have been taken to amend this for future analysis.

## Conclusion

The present study has included White and South Asian, and Black patient and control groups from East London, in an attempt to increase representation in research. We aimed to define the clinical phenotype of patients with PD from White, South Asian and Black ethnic groups. Our findings highlight that, despite an apparent younger age at assessment and similar PD duration and treatment burden, South Asian patients seem to have worse motor and cognitive phenotypes. A more severe motor and non-motor phenotype was also found in the Black patient group, although these patients were less well represented than South Asian in the present study. Further work is needed to understand if these differences arise as part of the natural course of the disease, or if socio-demographic factors lead to these differences.

## Supporting information

Supplementary References

## Data Availability

The datasets generated during and/or analyzed during the current study are available upon reasonable request from the corresponding authors.

## Author’s roles

AN designed the study.

KD, AZ, EC, TH, ABJ, JK recruited and assessed participants clinically.

AZ performed the analysis. AZ and KD wrote the paper.

AN, CM and SW reviewed the article. The rest of the authors also provided valuable feedback for the paper.

