## Supplementary Table 1, Supplementary Table 2, Supplementary Table 3, Supplementary Table 4 for "The East London Parkinson’s Disease Project – A case-control study in a diverse population"

**Supplementary information**

1. **Supplementary Tables**

|  | White n=52 | South Asian n=45 | Black n=8 | White vs South Asian | White vs Black | Black vs South Asian | Black vs South Asian vs Black |
| --- | --- | --- | --- | --- | --- | --- | --- |
|  |  |  |  | Uni | Uni | Uni | Uni |
|  |  |  |  | p, t(dof) | p, t | p, t(dof) | p, t(dof) |
| MERQ-PD-B Pesticides (n, %) | 2, 2% | 0, 0% | 0, 0% | **-** | **-** | **-** | 0.613^c^, 0.98 |
| MERQ-PD-B Head injury (n, %) | 9, 17% | 5, 11% | 1, 11% | **-** | **-** | **-** | 0.658^c^, 0.84 |
| MERQ-PD-B Caffeine previous consumption (n, %) | 51, 96% | 41, 91% | 9, 100% | **-** | **-** | **-** | 0.568^c^, 1.13 |
| MERQ-PD-B Caffeine previous consumption (mean ± SD) | 4.1 ± 2.3 | 3.1 ± 1.9 | 2.4 ± 0.8 | 0.026^e^, 1445.5 | 0.088^e^, 311 | 0.166^e^, 144.5 | **-** |
| MERQ-PD-B Caffeine current consumption (n, %) | 47, 89% | 37, 82% | 7, 78% | **-** | **-** | **-** | 0.547^c^, 1.21 |
| MERQ-PD-B Caffeine current consumption (mean ± SD) | 2.8 ± 2.3 | 2.1 ± 1.5 | 1.4 ± 1.2 | 0.331^a^, 0.946 | 0.101^e^, 302 | 0.166^e^, 0.63 | **-** |
| MERQ-PD-B Alcohol previous consumption (n, %) | 36, 75% | 7, 17.5% | 6, 67% | **-** | **-** | **-** | **<0.001^c^, 29.9** |
| MERQ-PD-B Alcohol current consumption (n, %) | 27, 56% | 1, 2% | 5, 56% | **-** | **-** | **-** | **<0.001^c^, 30.8** |
| MERQ-PD-B Smoking previous (n, %) | 24, 45% | 8, 18% | 0, 0% | **-** | **-** | **-** | **0.003^c^, 11.8** |
| MERQ-PD-B Smoking current (n, %) | 4, 8% | 1, 2% | 0, 0% | **-** | **-** | **-** | 0.479^c^, 1.47 |

Supplementary Table 1. Exposure in patients with Parkinson’s disease in the East London PD study. a - Kruskall Wallis test; c - Fisher's exact test; k - Logistic regression adjusted for gender; l - Logistic regression adjusted for gender, age, years of education. IMD - Index of Multiple Deprivation; SD - standard deviation; IQR - interquartile range; bold - p<0.004 (Bonferroni-adjusted for multiple comparisons).

|  | White n=96 | South Asian n=85 | Black n=24 | White vs South Asian | | White vs Black | | Black vs South Asian | |
| --- | --- | --- | --- | --- | --- | --- | --- | --- | --- |
|  |  |  |  | Uni | Multi | Uni | Multi | Uni | Multi |
|  |  |  |  | p, t(dof) | p | p, t | p | p, t(dof) | p |
| Smell test (median ± IQR) | 2 ± 1 | 2 ± 3 | 1 ± 1 | 0.911^e^, 872.5 | 0.917^f^, 0.986^g^ | 0.018^e^, 233.5 | 0.006^f^, 0.066^g^ | 0.037^e^, -1.17 | 0.006^f^, 0.119^g^ |
| HADS Depression (mean ± SD) | 6.3 ± 3.8 | 8.5 ± 5.2 | 6.8 ± 4.3 | 0.019^a^, 5.539 | 0.052^f^, **0.002^g^** | 0.627^e^, 560.5 | 0.055^f^, 0.435^g^ | 0.245^d^, 0.126(67) | 0.007^f^, 0.19^g^ |
| HADS Anxiety (mean ± SD) | 6.2 ± 4.3 | 7.9 ± 5 | 6.9 ± 4.5 | 0.077^a^, 3.128 | 0.152^f^, 0.026^g^ | 0.482^e^, 539.5 | 0.061^f^, 0.517^g^ | 0.514^e^, 385 | 0.021^f^, 0.281^g^ |
| RBDSQ (mean ± SD) | 3.9 ± 3.1 | 4.8 ± 3.1 | 5.9 ± 2.3 | 0.177^e^, 897 | 0.297^f^, 0.050^g^ | 0.035^e^, 166.5 | 0.831^f^, 0.013^g^ | 0.290^e^, 279.5 | 0.923^f^, 0.043^g^ |
| PSS2 (mean ± SD) | 17.1 ± 10.2 | 23.1 ± 11.7 | 19 ± 9.3 | 0.008^e^, 748 | 0.049^f^, **0.003^g^** | 0.464^e^, 295.5 | 0.177^f^, 0.495^g^ | 0.291^d^, -1.06(49) | 0.018^f^, 0.187^g^ |
| ESS (mean ± SD) | 8.7 ± 5.5 | 10.7 ± 7.1 | 9.3 ± 6.3 | 0.073^a^, 3.225 | 0.108^f^, 0.009^g^ | 0.743^e^, 987 | 0.137^f^, 0.595^g^ | 0.475^e^, 832 | 0.022^f^, 0.362^g^ |
| SCOPA-AUT (mean ± SD) | 18.1 ± 9.6 | 17.5 ± 10.7 | 16.9 ± 12.8 | 0.776^d^, 0.286(83) | 0.541^f^, 0.491^g^ | 0.456^e^, 250.5 | 0.263^f^, 0.654^g^ | 0.533^e^, 143.5 | 0.291^f^, 0.973^g^ |
| EQ5D5L VAS (mean ± SD) | 63.7 ± 16.7 | 53.3 ± 21.8 | 60.4 ± 22 | **0.001^e^, 5116** | 0.073^f^, 0.024^g^ | 0.740^e^, 1129.5 | **0.001^f^**, 0.335^g^ | 0.171^d^, 1.38(106) | 0.055^f^, 0.682^g^ |
| EQ5D5L index (mean ± SD) | 0.53 ± 0.26 | 0.38 ± 0.31 | 0.45 ± 0.33 | **<0.001^a^, 12.2** | 0.012^f^, 0.005^g^ | 0.359^e^, 1228 | **0.002^f^**, 0.144^g^ | 0.293^d^, 1.06(106) | 0.15^f^, 0.571^g^ |
| IMD decile (median ± IQR) | 4 ± 2 | 3 ± 1 | 3 ± 2 | 0.022^a^, 5.26 | 0.037^k^, 0.050^l^ | **0.003^a^, 8.73** | **<0.001^k^, 0.003^l^** | 0.101^e^, 611.5 | 0.023^k^, 0.016^l^ |

Supplementary Table 2. Clinical characteristics of patients with Parkinson’s disease in the East London Parkinson’s Disease project. d - two-sided T-test; e - Mann-Whitney U test; f - Logistic regression, adjusted for disease duration; g - Logistic regression, adjusted for disease duration, age and gender; k - Logistic regression, adjusted for gender; l - Logistic regression, adjusted for gender, age and years of education. SD - standard deviation; IQR - interquartile range; t - T statistic; dof - degrees of freedom; bold - p<0.004 (Bonferroni-adjusted for multiple comparisons).

| MoCA | | White n=91 | South Asian n=52 | Black n=16 | White vs South Asian | White vs Black | Black vs South Asian | Black vs South Asian vs Black |
| --- | --- | --- | --- | --- | --- | --- | --- | --- |
|  |  |  |  |  | Uni | Uni | Uni | Uni |
|  |  |  |  |  | p, t(dof) | p, t | p, t(dof) | p, t(dof) |
| PD normal cognition  (n, %) | MoCA > 25 | 50, 55% | 14, 27% | 4, 25% | **0.003^b^**, 11.5 | 0.084^c^, 4.95 | 0.688^c^, 0.75 | 0.006^c^, 14.45 |
| PD mild cognitive impairment (n, %) | MoCA 19-25 | 33, 36% | 27, 52% | 10, 63% |  |  |  |  |
| PD dementia (n, %) | MoCA < 19 | 8, 9% | 11, 21% | 2, 12% |  |  |  |  |
| PD normal cognition  (n, %) | MoCA > 25 | 50, 55% | 14, 27% | 4, 25% | **0.002^b^**, 9.4 | 0.027^c^, 4.88 | 0.879^c^,0.02 | **0.002^c^, 12.91** |
| PD cognitive impairment (n, %) | MoCA <= 25 | 41, 45% | 38, 73% | 12, 75% |  |  |  |  |

Supplementary Table 3. Cognitive impairment in patients with Parkinson’s Disease. b - Chi-squared test; c - Fisher's exact test; t - T statistic; dof - degrees of freedom; bold - p<0.004 (Bonferroni-adjusted for multiple comparisons).

| Chosen by highest UPDRS Left vs Right | | | | | | | | | |
| --- | --- | --- | --- | --- | --- | --- | --- | --- | --- |
| BRAINtest | **White**  **n=71** | **South Asian n=50** | **Black**  **n=13** | **White vs South Asian** | | **White vs Black** | | **Black vs South Asian** | |
|  |  |  |  | Uni | Multi | Uni | Multi | Uni | Multi |
|  |  |  |  | p, t(dof) | p | p, t | p | p, t(dof) | p |
| KS, n  (mean ± SD) | 36.5 ± 14.4 | 31.4 ± 12.5 | 32.2 ± 17.6 | 0.052^d^, 2(115) | 0.085^f^, 0.491^g^, 0.335^h^, 0.505^j^ | 0.358^d^, 0.9(80) | 0.007^f^, 0.348^g^, 0.317^h^, 0.385^j^ | 0.846^d^, 0.2(59) | 0.076^f^, 0.722^g^, 0.728^h^, 0.747^j^ |
| AT, ms  (mean ± SD) | 153.8 ± 70.2 | 204.9 ± 97.4 | 220.1 ± 93.1 | **0.002^a^, 7.4** | 0.171^f^, **<0.001^g^, <0.001^h^, <0.001^h^** | 0.018^e^, 261 | 0.697^f^, 0.008^g^, 0.012^h^, 0.008^j^ | 0.591^e^, 343 | 0.152^f^, 0.896^g^, 0.874^h^, 0.910^j^ |
| IS, ms^2^  (mean ± SD) | 18752.8 ± 19072.6 | 26332.1 ± 25727.1 | 22224.8 ± 13556.3 | 0.142^e^, 1008 | 0.306^f^, 0.070^g^, 0.075^h^, 0.062^j^ | 0.178^e^, 275 | 0.521^f^, 0.301^g^, 0.218^h^, 0.255^j^ | 0.72^e^, 257 | 0.214^f^, 0.854^g^, 0.957^h^, 0.871^j^ |
| Chosen by lowest KS Left vs Right | | | | | | | | | |
| BRAINtest | **PwP**  **n=145** | **HC**  **n=13** | | **p-value** |  |  |  |  |  |
| KS, n  (mean ± SD) | 34.7 ± 14.6 | 46.3 ± 8.8 | | **<0.001^i^**, 15.8(152) |  |  |  |  |  |
| AT, ms  (mean ± SD) | 180.3 ± 88.9 | 150 ± 38.6 | | 0.287^e^, 1081 |  |  |  |  |  |
| IS, ms^2^  (mean ± SD) | 22185.3 ± 22022.8 | 12145.7± 6590.8 | | 0.414^a^, 0.667 |  |  |  |  |  |

Supplementary Table 4. BRAINtest scores in the East London Parkinson’s Disease project. a - Kruskal Wallis test, d - two-sided T-test; e - Mann-Whitney U test; f - Logistic regression, adjusted for disease duration; g - Logistic regression, adjusted for disease duration, age and gender; h - Logistic regression, adjusted for disease duration, age, gender, and ON-OFF status, i - Welch’s ANNOVA test; j - Logistic regression, adjusted for disease duration, age, gender, and LEDD. KS - kinesia score, AT - akinaesia time; IS - incoordination score; SD - standard deviation; IQR - interquartile range; PwP – People with Parkinson’s; HC – healthy controls; t - T statistic; dof - degrees of freedom, ms - millisecond, cm - centimetre ; bold - p<0.004 (Bonferroni-adjusted for multiple comparisons).
