## Supplementary Figure 1, Supplementary Figure 2 for "The East London Parkinson’s Disease Project – A case-control study in a diverse population"

2. **Supplementary Figures**

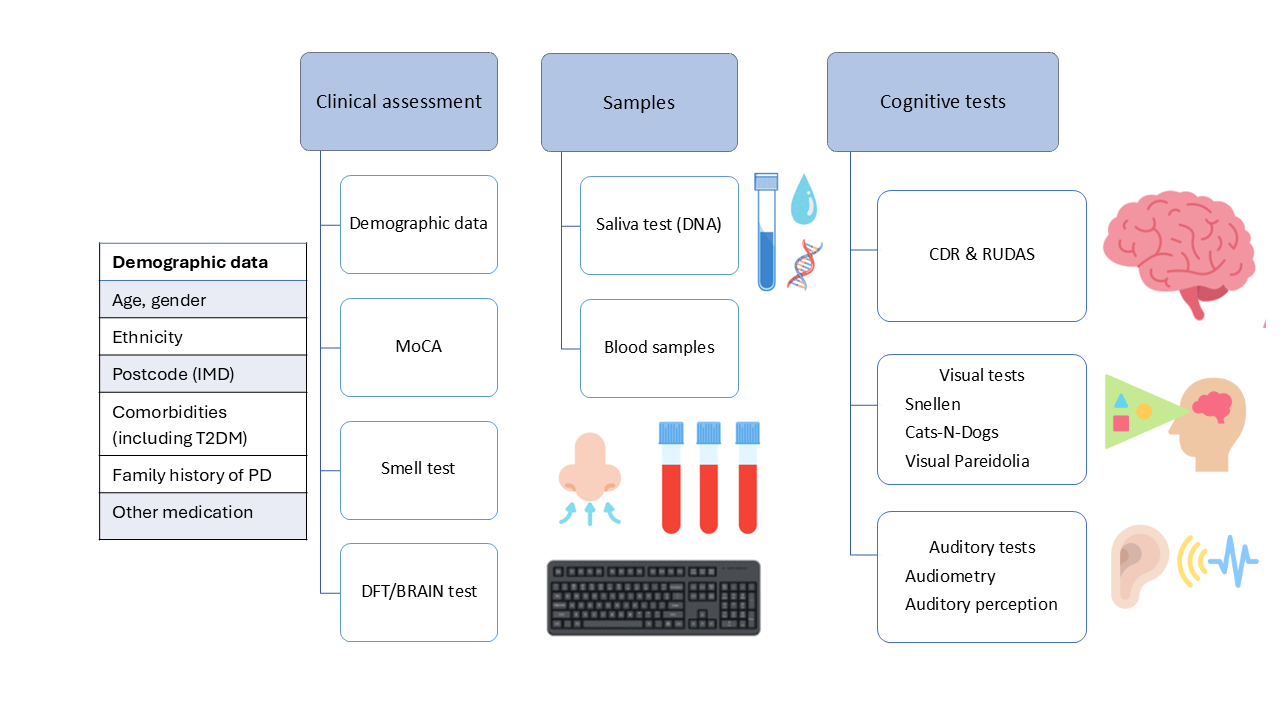

Supplementary Figure 1. Control visit protocol. CDR - Clinical Dementia Rating Scale^1^; IMD - Index of Multiple Deprivation^2^; RUDAS - Rowland Universal Dementia Assessment Scale^3^; Cats-N-Dogs - Cats and dogs test^4^.

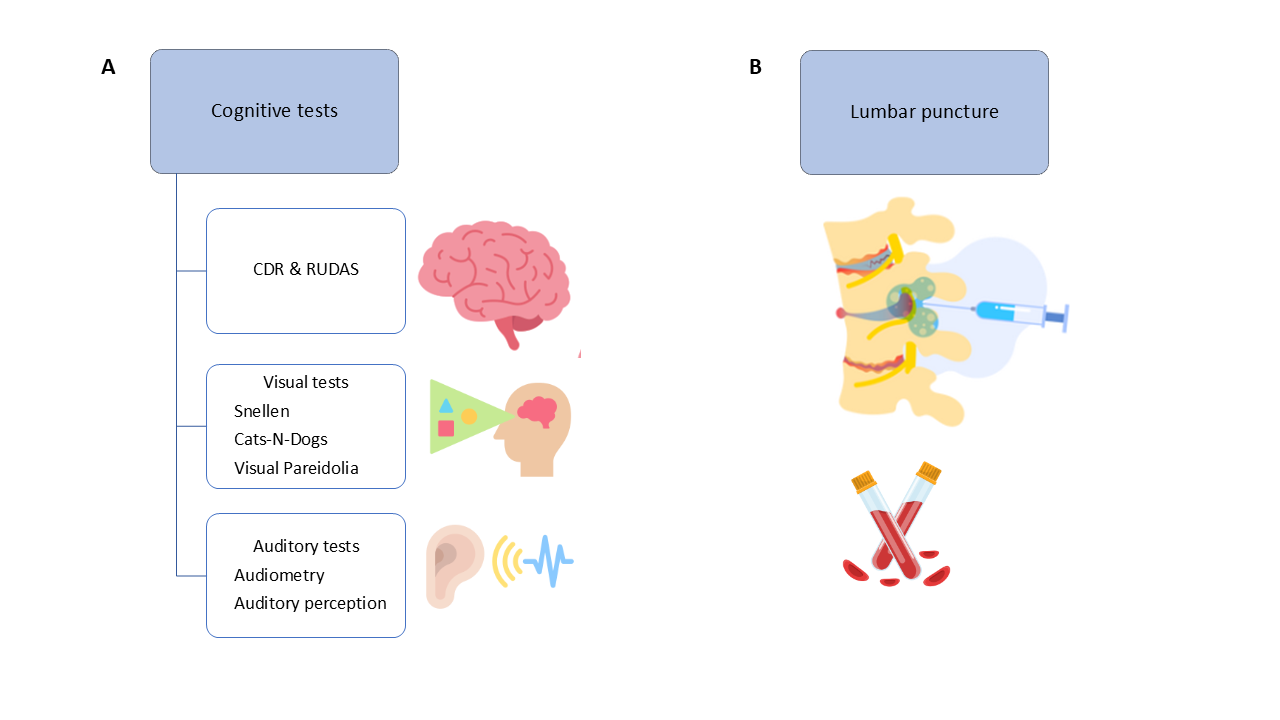

Supplementary Figure 2. A. Optional visit for the East London Parkinson’s Disease study. CDR - Clinical Dementia Rating Scale; RUDAS - Rowland Universal Dementia Assessment Scale; Cats-N-Dogs - Cats and dogs test. B. Optional visit for the East London Parkinson’s Disease Study consisting of a lumbar puncture.
