## Supplementary Appendix 1 for "The East London Parkinson’s Disease Project – A case-control study in a diverse population"

3. **Appendix 1**

From 218 patients, 6 South Asian and 1 Arabic individuals were unable to complete the assessment for MoCA. They were excluded from the cognitive analysis due to a language barrier at the outset (3) and other reasons (2 could not follow commands due to advanced Parkinson’s, and 2 had severe motor fluctuations). A total of 211 patients completed the MoCA assessment: 96 White, 79 South Asian, 24 Black patients and 12 from other backgrounds. Out of these 200 were performed in English, in some cases with a translator, and 15 directly in Bengali (validated translation).

A further 44 patients were excluded, as although they were able to complete part of the assessment, the results were unreliable due to: evident language barriers during tests (15, 7%), low levels of literacy (9, 4%), severe tremor (4, 2%), blindness (4, 2%), severe dyskinesia (1, 0.4%), severe dystonia (1, 0.4%) and other reasons (10, 5%). The majority of the patients who had unreliable scores were from South Asian backgrounds (24, 61%).

From 90 controls, 45 had MoCA assessments: 14 White, 30 South Asian, 1 other. Black controls did not have any available data. A further 6 scores in the controls were unreliable due to: language barrier (5, 11%) and other reasons (1, 2%).
