## Supplementary References for "The East London Parkinson’s Disease Project – A case-control study in a diverse population"

4. **Supplementary References:**
1. O’Bryant SE, Waring SC, Cullum CM, Hall J, Lacritz L, Massman PJ, et al. Staging dementia using Clinical Dementia Rating Scale Sum of Boxes scores: a Texas Alzheimer’s research consortium study. Arch Neurol. 2008 Aug;65(8):1091–5.
2. [Ministry of Housing, Communities, Local Government (2018 to 2021). English indices of deprivation 2019 [Internet]. GOV.UK; 2019 [cited 2024 Oct 23]. Available from:](http://paperpile.com/b/IXzDdP/rlYQ) <https://www.gov.uk/government/statistics/english-indices-of-deprivation-2019>
3. Storey JE, Rowland JTJ, Basic D, Conforti DA, Dickson HG. The Rowland Universal Dementia Assessment Scale (RUDAS): a multicultural cognitive assessment scale. Int Psychogeriatr. 2004 Mar;16(1):13–31.
4. Weil RS, Pappa K, Schade RN, Schrag AE, Bahrami B, Schwarzkopf DS, et al. The Cats-and-Dogs test: A tool to identify visuoperceptual deficits in Parkinson’s disease. Mov Disord. 2017 Dec;32(12):1789–90.
